## Supplementary material for "Impaired Estimated Glomerular Filtration Rate and Associated Factors among Adult Patients Living with HIV at Asella Referral and Teaching Hospital, Ethiopia: A Cross-Sectional Study": Supportive files

**Data Collection Tool**

**Title:** Impaired eGFR and Associated Factors among PLHIV at Asella Referral and Teaching Hospital, Ethiopia

**Part I: Socio-Demographic Characteristics**

| **No.** | **Question** | **Response Options** |
| --- | --- | --- |
| 1.1 | Sex | ☐ Male ☐ Female |
| 1.2 | Age | ☐ 18-39 years ☐ 40-65 years |
| 1.3 | Marital Status | ☐ Single ☐ Married ☐ Divorced ☐ Widowed |
| 1.4 | Highest Education Level | ☐ No formal education ☐ Primary (1-8 grade) ☐ Secondary (9-12 grade) ☐ Diploma/Degree+ |
| 1.5 | Ethnicity | ☐ Oromo ☐ Amhara ☐ Other: _____ |
| 1.6 | Religion | ☐ Muslim ☐ Orthodox ☐ Protestant ☐ Catholic ☐ Other: _____ |
| 1.7 | Occupation | ☐ Unemployed ☐ Government employee ☐ Self-employed ☐ Student ☐ Other: _____ |
| 1.8 | Residence | ☐ Urban ☐ Rural |
| 1.9 | Monthly Income (ETB) | ☐ <3,500 ETB ☐ ≥3,500 ETB |
| 1.10 | Family History of Kidney Disease | ☐ Yes ☐ No |

**Part II: Behavioral Factors**

| **No.** | **Question** | **Response Options** |
| --- | --- | --- |
| 2.1 | Smoking Status | ☐ Never smoked ☐ Former smoker ☐ Current smoker |
| 2.2 | Smoking Frequency (if current smoker) | ☐ Monthly ☐ Weekly ☐ Daily |
| 2.3 | Alcohol Consumption | ☐ Never ☐ Occasionally ☐ Weekly |

**Part III: HIV-Related & Clinical Factors**

| **No.** | **Question** | **Response Options** |
| --- | --- | --- |
| 3.1 | CD4 Count (cells/µL) | ☐ <200 ☐ ≥200 |
| 3.2 | Blood Pressure (mmHg) | ______ / ______ mmHg |
| 3.3 | Weight (kg) | ______ kg |
| 3.4 | Height (m) | ______ m |
| 3.5 | BMI (kg/m²) | ______ kg/m² |
| 3.6 | WHO Clinical Stage | ☐ Stage I ☐ Stage II ☐ Stage III ☐ Stage IV |
| 3.7 | Opportunistic Infections | ☐ Yes ☐ No |
| 3.8 | Comorbidities | Diabetes: ☐ Yes ☐ No Hypertension: ☐ Yes ☐ No |
| 3.9 | ART Regimen | ☐ 1st-line ☐ 2nd-line ☐ 3rd-line |
| 3.10 | Tenofovir-Based Regimen | ☐ Yes ☐ No |
| 3.11 | ART Adherence | ☐ No interruptions ☐ History of interruptions |
| 3.12 | Renal Function | Serum Creatinine: ______ mg/dL eGFR (CKD-EPI 2021): ______ mL/min/1.73m² |
| 3.13 | Functional Status | ☐ Working ☐ Ambulatory |
